# Balancing Relapse Risk and Agency in Buprenorphine-naloxone Treatment: A Qualitative Needs Assessment to Inform Patient-Centered Care

**DOI:** 10.64898/2026.08.21.26360804

**Authors:** Thomas J. Reese, Mauli V. Shah, Adam Wright, Michael Matheny, David Marcovitz, Kristopher Kast, John F P Bridges, Hilary A. Tindle, Amanda von Horn, Carolyn Audet

## Abstract

**Objectives:** Outpatient buprenorphine-naltrexone (bup-nx) treatment reduces overdose risk, yet many patients still return to use or disengage from treatment. We sought to understand how patients and prescribers experience and manage relapse risk, monitoring, and treatment agency in routine bup-nx treatment to identify gaps in current practice.

**Methods:** We conducted a qualitative needs assessment using semi-structured, critical-incident interviews with patients receiving outpatient bup-nx and prescribers who manage bup-nx treatment. Interviews examined situations involving relapse risk and empowerment in treatment decisions. We structured data collection and analysis using the Theoretical Domains Framework and COM-B model to characterize determinants. Transcripts were coded deductively and inductively until code-level saturation was reached.

**Results:** Participants (9 patients, 8 prescribers) described nine treatment needs mapped to the Capability, Opportunity, and Motivation components of the COM-B model. These themes highlighted how patient agency in bup-nx treatment was constrained by physiologic and emotional states, with withdrawal, craving, pain, and distress often overriding longer-term goals. Relapse vulnerability was experienced as dynamic and intensifying between visits, while clinical detection remained anchored to visit-bound assessments, urine drug testing, refill patterns, and crisis-driven contact, creating blind spots. Structural friction (pharmacy rules, insurance disruptions, transportation and housing instability), stigma from family and recovery communities, and motivational processes tied to fluctuating readiness and trust in monitoring further shaped engagement, disclosure, and dosing decisions; the same monitoring tools could either support honest disclosure or provoke concealment when perceived as punitive.

**Conclusions:** Relapse risk and agency in bup-nx treatment are negotiated as dynamic processes within structurally constrained and trust-sensitive systems. Addressing the identified capability, opportunity, and motivation gaps will require patient-centered, trust-preserving approaches to monitoring and shared decision-making.

## INTRODUCTION

Opioid use disorder (OUD) remains a leading cause of preventable morbidity and mortality in the United States, despite expanded access to evidence-based treatment.^1,2^ Buprenorphine-naloxone (bup-nx) improves treatment retention and substantially reduces overdose risk when compared with no medication, yet many patients still return to use or disengage, particularly early in treatment and after transitions in care intensity.^3–5^ Improving outcomes, therefore, requires attention not only to initiating bup-nx, but also to how return-to-use risk is monitored and managed over time in routine outpatient care.^6,7^

Among the three approved medications for OUD, bup nx is the most commonly used in office based care and is frequently embedded in treatment systems that condition medication access on monitoring and behavioral contingencies. As a result, questions of how evolving relapse risk is assessed and managed are especially pronounced in bup nx treatment. Relapse vulnerability during bup-nx treatment is dynamic rather than static, with risk changing over weeks to months rather than remaining fixed.^8,9^ Patient risk often fluctuates between visits in response to physiological constraints such as withdrawal, craving, and pain, affective states including stress, trauma, and co occurring mental illness, and changes in social context, housing, or relationships.^9,10^ In contrast, most bup-nx care is still organized around visit assessments and retrospective indicators such as urine drug testing, prescription refill patterns, or crisis driven encounters, rather than continuous monitoring of evolving risk.^11–13^ This structural mismatch forces clinicians to balance relapse, overdose, and diversion concerns against patient agency and preferences using incomplete or delayed information.^11,14^ Prior work on patient centered care and medications for OUD has described barriers to initiation, retention, and shared decision making, yet it offers limited insight to how patients and clinicians experience and navigate this tension in the context of dynamic, between visit relapse risk.^11,12^

Understanding this tension requires methods that can capture the interplay of physiologic dependence, emotional vulnerability, stigma, and social environment in shaping relapse risk and treatment decisions.^15,16^ Qualitative inquiry is well suited to explain how patients and clinicians interpret and respond to evolving risk in everyday practice, including when monitoring is experienced as supportive versus punitive.^17–19^ The Theoretical Domains Framework (TDF) provides a comprehensive set of behavioral determinants that can be used to structure data collection and analysis in ways that inform intervention development, and mapping these determinants onto the COM-B model (Capability, Opportunity, Motivation–Behavior) allows synthesis at a higher level, clarifying where barriers and enablers to safe, patient-centered bup-nx care cluster within the broader behavioral system.^20,21^

We sought to understand how patients and prescribers experience and manage relapse risk, monitoring, and treatment agency in routine bup-nx care and to identify resulting gaps in current practice. Taken as a qualitative needs assessment, these theory-informed findings highlight capability, opportunity, and motivation gaps that help explain why relapse risk, disengagement, and tensions around agency persist despite access to effective medication. These insights help identify targets for designing patient-centered strategies to support safer, more collaborative bup-nx treatment.

## METHODS

### Study Design

In this qualitative study, we used semi-structured interviews to examine how patients and prescribers experience and manage relapse risk, monitoring, and treatment agency in routine bup-nx care.^22,23^ We used the TDF to structure data collection and analysis, and findings were synthesized using the COM-B model to support a qualitative needs assessment focused on practice gaps relevant to intervention development.^24–29^ This qualitative study is reported in accordance with the Standards for Reporting Qualitative Research and it was approved by Vanderbilt University Medical Center Institutional Review Board.^30^

### Setting and Participants

The study took place within Vanderbilt University Medical Center, a large health system in the Southeast United States that offers comprehensive addiction services across inpatient and outpatient settings. These services include inpatient addiction treatment, transitional care primarily for discharged patients (Vanderbilt Bridge Clinic), and longitudinal care for addiction and co-occurring disorders (Vanderbilt Recovery Clinics).^17,31,32^ These services care for patients with OUD, supported by multidisciplinary care teams comprising physician and nurse practitioner specialists in addiction psychiatry, internal medicine, infectious diseases, and pain-anesthesia, along with licensed social workers, nurse case managers, and recovery coaches. The care teams have adopted a patient-centered approach, tailoring holistic treatment plans to the unique needs of each patient.^33^ They build trust through empathy and understanding to empower patients while navigating their recovery journey. The overarching goal of treatment is to initiate patients on medication for OUD during hospitalization, stabilize them post-discharge, and transition them to long-term treatment programs over the course of a year or more. The combination of longitudinal addiction treatment and patient-centered approaches makes this an ideal setting for this study.

We purposively sampled prescribers and patients involved in outpatient bup-nx treatment. Prescribers were eligible if they were responsible for managing patients with OUD including those treated with bup-nx and treating co-occurring substance use and mental health conditions. To capture diverse perspectives, we sampled prescribers across levels of training and clinical effort in addiction services (e.g., attending physicians, fellows, and residents). Prescribers were invited to participate by a clinician-investigator who practices in the addiction clinics (DM). Eligible patients were currently prescribed bup-nx in an outpatient addiction clinic. We purposively sampled patients based on phase of treatment (e.g., early vs. stable), presence of co-occurring psychiatric conditions, and perceived stability to capture variation in relapse risk and decision-making contexts. Patients were invited to participate by their prescriber following a scheduled visit. All participants provided informed consent and received a $50 gift card as compensation for their time.

### Data Collection

We used the Critical Incident Technique to elicit detailed narratives about specific high-salience events related to relapse risk and empowerment in treatment decisions.^22^ This approach is well suited to health services research questions examining how particular experiences and behaviors contribute to care delivery, including when processes succeed or break down.^23,34,35^ The interview guide was developed and piloted by two researchers with experience in qualitative methods (TJR and CA) and intervention design, and it was organized around TDF domains to ensure coverage of relevant behavioral determinants.^17,36–39^ We developed questions focusing on two incidents: 1) a situation involving risk of relapse and 2) situation involving empowerment to make important treatment decisions. Prescribers were asked to recall a patient who relapsed or was at high risk for relapse and a different patient whom they felt they had empowered to make a key treatment decision (e.g., dose adjustment, formulation change, or visit frequency). Patients were asked to recall a time when they relapsed or felt at high risk for relapse and a time when a prescriber empowered them to make an important treatment decision. For each incident, we prompted participants to describe what led up to the event, what happened during the encounter, and what followed, with particular attention to factors that helped or hindered the situation. Probing questions were mapped to TDF domains (e.g., knowledge, beliefs about consequences, social influences, environmental context) to elicit determinants of behavior relevant to relapse prevention and agency. Interviews were conducted by one author (TJR), with a second author present to take notes and ask clarifying questions as needed (MS). Field notes were used to document initial impressions about context, nonverbal cues, and potential analytic leads. Interviews were conducted from September 2024 through August 2025.

### Data Analysis

We planned an initial sample of 15–25 interviews, in line with methodological work indicating that code– or theme-level saturation in relatively narrow, theory-informed interview studies is typically reached with fewer than 25 participants.^40–42^ Transcripts were coded both sequentially and in parallel, allowing the team to monitor the emergence of new codes and to judge when code saturation had been achieved, defined as the point at which no new codes were identified in successive interviews. We used both inductive and deductive approaches anchored in the TDF and COM-B frameworks. The TDF, developed from a synthesis of psychological theories, provides 14 domains capturing psychological, social, and environmental influences on behavior and is widely used to diagnose implementation-relevant barriers and facilitators.^39^ The COM-B model conceptualizes behavior as a function of capability, opportunity, and motivation and offers a higher-level structure for organizing determinants into actionable targets.^29^ We began with a deductive coding framework based on the TDF, developing an initial codebook from the TDF domains and the interview guide that was iteratively refined during early coding. Two authors applied TDF codes to all transcripts (TJR and MS), with regular meetings and an additional author (CA) to review excerpts, resolve disagreements, and refine code definitions. Following TDF coding, we mapped coded segments to COM-B components (capability, opportunity, motivation) to synthesize findings at a broader behavioral-system level and to compare patterns across patients and prescribers.

Within each COM-B component, we then used an inductive approach to identify themes that captured how relapse risk, monitoring, and treatment agency were experienced and managed in practice. This involved open coding within each COM-B category, grouping related codes into candidate themes, and refining themes based on their ability to explain the observed trade-offs between minimizing relapse risk and supporting patient agency, as well as their relevance for potential intervention design.^29^ Throughout analyses, we attended to tensions that could not be fully resolved (e.g., safety vs. agency and standardization vs. individualization) and to differences and alignments between patient and prescriber perspectives. We documented analytic decisions in memos and discussed emerging interpretations in team meetings to enhance rigor and reflexivity. COM B was used as an integrative lens rather than a primary coding schema, enabling translation of detailed qualitative findings into a coherent behavioral assessment of gaps in routine bup-nx care. Coding was conducted iteratively across interviews, and no additional codes were identified in the final interviews, indicating that code level thematic saturation had been approached.

## RESULTS

We completed interviews with 9 patients and 8 prescribers (**Table 1**). Using the COM B model as a high level structure, we identified nine themes mapped to Capability, Opportunity, and Motivation (**Figure 1**) describing how patients and prescribers experienced relapse risk, monitoring, and treatment decision making in routine bup nx care. Across domains, participants consistently described a mismatch between continuous, evolving relapse risk and visit-bound care processes, shaping how agency, safety, and trust were negotiated in practice.

**Figure 1.**
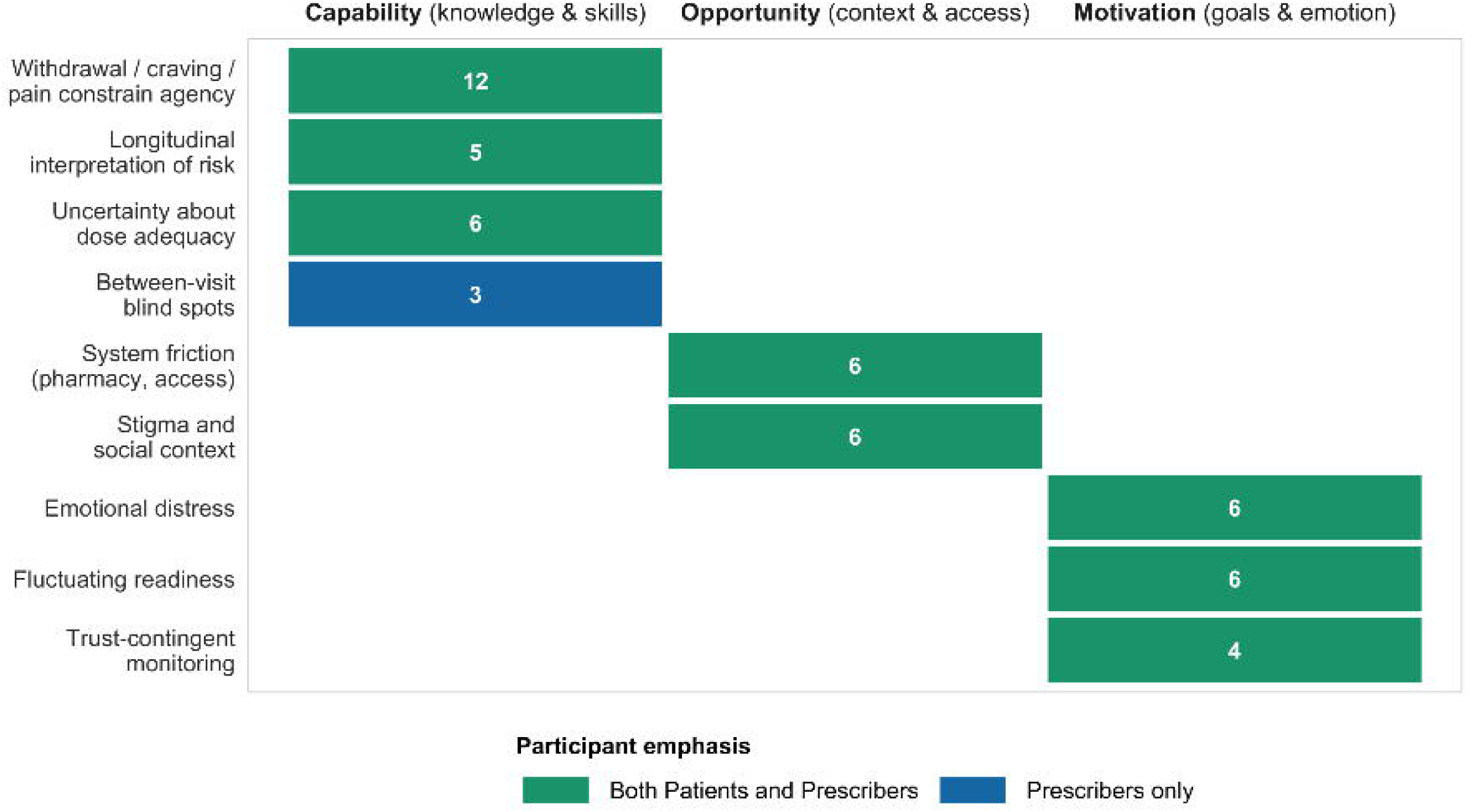
Heat map of buprenorphine treatment needs by themes and COM-B domains. Colors indicate whether themes were emphasized by prescribers only or by both patients and prescribers; no themes were identified by patients alone. Numbers show total coded quotes. Rows show key analytic themes, and columns show COM B domains (Capability, Opportunity, and Motivation) based on coded patient and prescriber interviews.

**Table 1.** Participant characteristics.

| **Table 1. Participant characteristics.** |  |  |
| --- | --- | --- |
| **Characteristic** | **Patients (N = 9)** | **Prescribers (N = 8)** |
| Age, mean (SD), years | 49 (10.4) | — |
| Female, n (%) | 3 (33) | 3 (38) |
| Racial/ethnic minority, n (%) | 1 (11) | — |
| Employed, n (%) | 3 (33) | — |
| Housed (stable housing), n (%) | 8 (89) | — |
| Co-occurring mental health diagnosis, n (%) | 7 (78) | — |
| Attending (versus trainee), n (%) | — | 4 (50) |

### Constraints on Choice and Risk Interpretation (Capability)

Providers and patients described decision-making about bup-nx as driven by acute physiological states and longitudinal interpretation rather than stable preferences or abstract goals.

#### Bounded agency driven by withdrawal, craving, and pain

Patients consistently described withdrawal, craving, and pain as dominant drivers of behavior that constrained perceived choice. Fear of withdrawal was often described in extreme terms and frequently outweighed longer-term goals such as tapering or reducing medication. Providers similarly emphasized that inadequate treatment of craving, rather than visible withdrawal alone, often preceded relapse, particularly in the fentanyl era and in the context of comorbid pain.

> *“Once that feeling comes – if I ain’t got it, I’m going to get whatever I can to make me feel better. I’d rather be dead than feel that feeling. I can’t go through it.”* (Patient 8)
>
> *“People can get enough buprenorphine to not be sick, but they’re still craving – and that’s where relapse happens.”* (Provider 2)
>
> *“A lot can change before your next visit.”* (Patient 3)

Together, these accounts portray agency in bup-nx treatment as conditional under strong physiological constraint rather than as a free or stable preference. Patients described withdrawal, craving, and pain as overpowering states that narrowed perceived choice and shifted priorities toward immediate relief. Fear of withdrawal was often described as eclipsing longer-term goals such as tapering or dose reduction. Clinicians likewise noted that patients could appear stable during visits while still experiencing persistent craving that escalated risk outside clinical encounters. Participants emphasized that these constraints often intensified between visits, when symptoms, stressors, or access barriers could worsen without real-time clinical visibility into emerging vulnerability.

#### Risk assessment as a longitudinal, interpretive process

Prescribers characterized relapse risk assessment as an interpretive process based on patterns over time rather than discrete indicators. Missed visits, refill timing concerns, and behavioral changes were interpreted differently depending on prior stability and life context. Patients echoed this emphasis on early, subjective warning signs (e.g., stress, fear, and exposure to triggers) that often precede observable relapse.

> *“Knowing the person matters – missed appointments mean different things for different people.”* (Provider 2)
>
> *“I usually know when I’m heading the wrong way. It starts with stress.”* (Patient 3)
>
> *“I have to monitor myself all the time really… I’m just aware that the potential’s always there to relapse.”* (Patient 5)

Together, these narratives suggest that relapse vulnerability was experienced by patients as dynamic and continuously self monitored, while clinical awareness often relied on retrospective, visit based signals, reinforcing episodic interpretation rather than continuous shared awareness.

#### Knowledge gaps shaping medication decisions

Patients described uncertainty about dosing, duration of treatment, and what “doing well” on bup-nx meant.

Several hesitated to request dose changes or framed interest in tapering as a marker of success. Providers viewed these assumptions as common and frequently misaligned with evidence regarding relapse risk and retention.

> *“I don’t know if I’m at the maximum dose – I’ve never asked.”* (Patient 2)
>
> *“Wanting to come off [bup-nx treatment] doesn’t always mean someone is ready.”* (Provider 8)

Across accounts, medication decisions were often shaped by uncertainty and assumptions rather than proactive, collaborative adjustment, reflecting a lack of shared, timely information about symptoms and risk.

### Between-Visit Blind Spots and System Friction (Opportunity)

Participants identified structural features of outpatient care that limited visibility into emerging relapse risk, particularly between scheduled encounters and during transitions in care intensity.

#### Relapse risk emerging between scheduled encounters

Both patients and prescribers emphasized that relapse vulnerability often emerged or became more difficult to manage between visits, particularly during transitions to less frequent contact. Providers described periods of heightened disengagement when visit intervals were lengthened (e.g., biweekly to monthly), while patients emphasized that meaningful changes in stress, symptoms, or exposure to triggers could occur at any point, often well before the next appointment. Rather than risk steadily accumulating between visits, participants described vulnerability as episodic and change driven, with clinic visits serving as intermittent anchors within otherwise unstable contexts.

> *“Going from every two weeks to monthly – that’s when people disappear.”* (Provider 1)
>
> *“A lot can change before your next visit.”* (Patient 3)

Taken together, these accounts suggest that relapse risk was shaped less by calendar time alone than by rapid, unpredictable changes in patients’ lives, creating blind spots in an outpatient care model organized around scheduled encounters.

#### System friction compounding vulnerability

Patients described pharmacy rules, refill timing, transportation barriers, housing instability, and insurance constraints as stressors that amplified relapse risk. Providers similarly identified access disruptions as high-risk “loss points” in care.

> *“When the pharmacy says come back tomorrow, that’s dangerous.”* (Patient 3)
>
> *“We lose people when access [to bup-nx] gets shaky.”* (Provider 6)

In response to these access uncertainties, some patients described adjusting or conserving their bup-nx doses to avoid running out, framing this as a protective response to fear of withdrawal rather than misuse. These strategies were shaped by prior experiences with refill delays and pharmacy rules and varied by trust: patients with established relationships described greater prescribing flexibility, while others encountered rigid policies that heightened vulnerability between visits. Collectively, these accounts highlight how structural and logistical barriers often interacted with physiological and emotional stressors, increasing vulnerability outside the clinic’s immediate awareness.

#### Social context shaping engagement

Patients described stigma from family, recovery communities, and social networks that influenced how they viewed bup-nx treatment. Some questioned the legitimacy of their recovery based on others’ feedback.

Providers noted that stigma could contribute to under-treatment or reluctance to adjust medication.

> *“Some people don’t think Suboxone counts [as quitting opioids].”* (Patient 9)
>
> *“Internalized stigma keeps people on lower doses than they need.”* (Provider 6)

These narratives suggest that social context shaped engagement, disclosure, and willingness to adjust treatment in ways that were not consistently visible during routine clinical encounters.

### Emotion, Readiness, and Trust in Monitoring (Motivation)

Motivational processes were closely tied to emotional distress, perceived readiness, and trust in monitoring practices, shaping whether risk was disclosed early or concealed until crisis.

#### Emotional distress as an early driver of relapse vulnerability

Patients frequently described fear, trauma, grief, and hopelessness as precursors to relapse risk. Providers identified emotional distress as an early warning signal that often precedes substance use or treatment disengagement.

> *“Fear [often related to withdrawal] was the main reason I relapsed.”* (Patient 2)
>
> *“When hope drops, risk goes up fast.”* (Provider 4)

Across interviews, emotional vulnerability was described as an important antecedent to observable behavioral change, yet it was not systematically visible within visit-based care.

#### Readiness and motivation as fluctuating states

Both patients and providers emphasized that engagement depended on internal readiness. Providers described harm-reduction approaches that avoided coercion, while patients stressed that recovery required personal willingness.

> *“You can’t force someone to change.”* (Provider 4)
>
> *“You have to want it yourself.”* (Patient 7)

These accounts depict readiness and motivation as fluctuating states, suggesting that fixed expectations for monitoring or escalation can be misaligned with patients’ evolving goals and may contribute to disengagement when applied inflexibly.

#### Monitoring practices as trust-contingent

Participants consistently described monitoring as either supportive or threatening depending on how it was enacted. Fear of punitive consequences led some patients to delay disclosure of cravings or substance use. Providers emphasized that transparency and relationship quality shaped whether monitoring facilitated honesty.

> *“If it feels like punishment, trust disappears.”* (Provider 1)
>
> *“I worry that if I’m honest, I’ll lose my meds.”* (Patient 9)

Overall, monitoring practices were experienced as trust-contingent rather than inherently supportive or coercive; the same tools could either strengthen or undermine disclosure depending on framing and use.

### Summary

Across capability, opportunity, and motivation domains, participants described a common underlying problem: relapse risk was experienced as continuous and evolving, while outpatient bup-nx treatment remained organized around periodic encounters. In this context, clinicians often became aware of escalating risk only after missed visits, crises, or disengagement, while patients managed vulnerability largely on their own between appointments. Agency and safety were therefore negotiated under conditions of incomplete and delayed information, in which trust in monitoring shaped whether risk signals were shared or concealed. Together, these findings highlight persistent gaps in how relapse risk and empowerment are navigated in routine bup-nx treatment and point to the need for more systematic, trust preserving approaches to monitoring and shared decision making that are better able to respond to rapid, between visit changes in risk, without requiring continuous, high intensity surveillance.

## DISCUSSION

We found that patients and prescribers experienced a consistent mismatch between dynamic, evolving relapse risk and outpatient bup-nx treatment organized around periodic encounters. Across interviews, relapse vulnerability was described as fluctuating between visits, while clinical detection of risk relied largely on scheduled appointments, urine drug screening, and crisis-driven contact. In this setting, agency, safety, and trust were negotiated under conditions of incomplete and delayed information.

A central finding was that patients’ agency in bup nx treatment was shaped and often constrained by physiologic and emotional states, not just by their interactions with clinicians.^43^ Patients described withdrawal, craving, and pain as powerful drivers that limited perceived choice, while clinicians noted that persistent craving frequently preceded relapse even when overt withdrawal was controlled.^44–46^ Participants also described uncertainty about dose adequacy, treatment duration, and what it meant to be “doing well” on bup-nx, echoing prior work showing that subtherapeutic dosing and unclear treatment goals are associated with higher craving, earlier dropout, and relapse. Together, these findings suggest that challenges in decision making reflect not simply motivation or adherence, but a capability gap in shared understanding of symptoms, risk, and appropriate treatment adjustment.^43^

Participants also highlighted substantial between-visit blind spots. Missed appointments, refill urgency, changes in visit cadence, and pharmacy or insurance disruptions were described as high-risk junctures that were difficult to interpret and often became visible only after deterioration or disengagement.^47^ Social context further shaped engagement: stigma from family, peers, providers, or recovery communities influenced whether patients disclosed symptoms, accepted higher doses, or viewed bup-nx and other medications for OUD as legitimate treatment.^48–50^ These findings indicate that opportunity-related barriers in bup-nx care extend beyond clinic visits themselves and include the broader structural and social environment in which treatment is managed.^47,51^

Motivational processes were closely tied to emotional distress, readiness, and trust in monitoring practices. Patients described fear, grief, trauma, and hopelessness as early drivers of relapse vulnerability, while clinicians recognized that these states often preceded overt substance use or missed visits.^52^ At the same time, both groups emphasized that engagement depends on internal readiness and that change cannot be forced, consistent with motivational interviewing principles that position lasting change as emerging from patients’ own motivation rather than external pressure.^53^ Monitoring was experienced as trust-contingent: the same practices could support honesty when framed collaboratively or provoke concealment and relationship rupture when perceived as punitive.^54,55^ Fear that disclosure could jeopardize medication access appeared to delay reporting of cravings, lapses, and side effects, as described in prior work where patients worried that abnormal urine drug tests or increased monitoring might threaten their bup-nx treatment.^54,56^

These findings have direct implications for intervention design. They suggest that, although participants did not name “measurement-based care,” they consistently endorsed its core elements: routine self-monitoring, structured feedback, shared interpretation, and proportionate responses to emerging risk.^57^ Patients wanted simple, non-punitive ways to see when they were doing well versus when they were vulnerable and to raise concerns safely, while clinicians wanted concise, workflow-compatible information to guide dosing, visit frequency, and escalation decisions.^58^ Participants also emphasized that any monitoring strategy must remain low-burden, trust-preserving, and individualized rather than algorithmic. Together, these points provide both a rationale and concrete design requirements for the measurement-based care strategy described in the companion intervention paper.

This study has several strengths, including inclusion of both patient and prescriber perspectives, use of established behavioral frameworks, and focus on concrete critical incidents rather than abstract attitudes. Limitations should also be noted. The study was conducted in an academic addiction psychiatry setting with an explicit patient-centered orientation, which may limit transferability to other treatment models. In addition, patients who had discontinued bup-nx or never initiated treatment were not represented. Because this was a qualitative study, the findings are not statistically generalizable but may be useful in settings where relapse risk, agency, and monitoring are negotiated in similar ways.

## CONCLUSION

Relapse risk and agency in bup-nx treatment emerged not as fixed characteristics assessed during isolated visits, but as dynamic processes unfolding between encounters within structurally constrained systems.

Patients and clinicians described care that often struggles to keep pace with fluctuating vulnerability, limited shared visibility, and fragile trust around monitoring. Addressing these needs will likely require approaches that improve timely, non-punitive visibility into risk and support collaborative, proportionate responses, thereby strengthening patient-centered bup-nx treatment.

## Data Availability

All data produced in the present study are available upon reasonable request to the authors and subject to data sharing requirements by the institution.

## ACKNOWLEDGEMENTS

Primary funding for this study was provided by the Agency for Healthcare Research and Quality (AHRQ) through grant K08HS029695 (TJR).

---

**Supplement Figure 1.** Patient interview guide.

## Notes

### Competing Interest Statement

The authors have declared no competing interest.

### Author Declarations

Vanderbilt University Medical Center Institutional Review Board gave ethical approval for this work.

