## Supplement Figure 1 for "Balancing Relapse Risk and Agency in Buprenorphine-naloxone Treatment: A Qualitative Needs Assessment to Inform Patient-Centered Care"

Subject ID #: ___________________________________
Interviewer: ___________________________
Date of Interview: __ __ / __ __ / __ __ __ __
MM DD YYYY
Start Time: __ __ : __ __ AM / PM

**<u>Introduction</u>**

Hello, my name is ____________ and I am a member of a team from Vanderbilt University Medical Center that is working to better understand how to improve treatment of opioid use disorder.

Thank you for taking the time to speak with me about your experiences and thoughts. Your participation in this interview is voluntary, anonymous, and confidential. The interview is expected to take about 60 minutes. No one outside of the project team will know your responses. You may choose not to answer a specific question during this interview. Also, if you no longer want to speak with me, just let me know and we can end the interview.

The primary risk for you participating in this interview is that you may be uncomfortable talking with me about this topic.

So that I can make sure we get the information correct, is it okay with you that we record our conversation? I want to make sure we don't miss any important details, and I will not use your name or personal information in any reports from today's discussion. I will use code number, so your name is not written down or recorded. I will keep the recording confidential on a secure network so no one outside of the project team will have access to it.

You are the expert, what I would like to do is understand how to better support recovery for patients between office visits. So, thank you so much for helping me. While we are talking about past and future care, **this conversation will not impact your care in any way**. I am asking you these questions so that we can create **better communication between patients and their providers** when it comes to treatment for opioid use disorder.

My phone number and email are on the contact information we have provided, so call or email me if you have any questions afterward. Do you have any questions? [Yes/No] Okay, let's begin.

**<u>Opening questions:</u>**

1. Can you tell me about how long you had been using opioids?
   a. **Probe:** still use opioids, how often
2. What other substances, if any, do you use (e.g., alcohol, stimulants)?
3. How would you define relapse?
4. Can you tell me about a time during treatment when you felt like you were going to relapse?
   a. **Probes:** What led up to the feelings of relapse (e.g., opioid use/withdrawal, cravings, lack of medication)? What did you do?

**<u>Questions about avoiding relapse and using buprenorphine:</u>**

5. Can you tell me about what being successful in treatment looks like for you? **(Motivation – Goals, Optimism**)
   a. **Probes:** avoiding relapse, relationships, normal daily activities, employment, length of treatment
6. Can you tell me about how you plan to achieve the success you mentioned? (**Motivation – Beliefs about capabilities, Intentions**)
   b. **Probes:** staying in treatment, taking medications
7. For patients like you, what do you think are the biggest challenges to staying in treatment? (**Motivation – Social/professional role and identity; Opportunity – Environmental context and resources; Opportunity – Social Influences**)
   a. **Probes:** getting prescriptions for buprenorphine, people close to you, financial resources, stable living environment, time for treatment, stigma
8. How do you think buprenorphine plays a role in your recovery? (**Capability – Knowledge**)
9. What do you think are the best or worst parts about taking buprenorphine? (**Motivation – Emotion, Reinforcement; Capability – Memory, attention, and decision processes**)
   a. **Probes:** what's difficult e.g., remembering, cravings, withdrawals, mood, sedation

10. How do you think skipping doses or stopping treatment early will affect your recovery? (**Motivation – Beliefs about consequences**)
11. Can you tell me about any changes <u>you</u> have made in taking buprenorphine that helps? (**Capability – Skills and Behavioral regulation**)

**<u>Questions about participating in treatment decisions:</u>**

12. Can you tell me about a time when you reflected on your treatment, such as urine analysis results or medication use and were encouraged (or discouraged) to make decisions with your provider.
    a. **Probes:** What led up to the feelings of encouragement/discouragement? What happened in the end?
13. Can you tell me how you would like to participate in treatment decisions? (**Motivation – Goals**)
14. How do you think discussing treatment decisions with your provider helps? (**Capability –Knowledge; Motivation – Reinforcement; Motivation – Optimism**)
    a. **Probes:** primary benefits
15. Can you tell me what you think your role is in assessing risk for relapse and helping with treatment decisions? (**Motivation – Social/professional role and identity**)
16. Can you tell me how you currently monitor your symptoms and make changes that help? (**Capability – Behavioral regulation**)
17. Can you tell me how you have communicated with your provider about treatment needs (**Capability – Skills; Capability –Memory, attention, and decision processes; Motivation – Intentions; Motivation – Beliefs about capabilities**)
    a. **Probes:** what was difficult about the conversation? did you remember to discuss everything that concerns you? Do you intend to do this in the future?
18. How do you think it would change your recovery if you don't routinely assess your treatment? (**Motivation – Beliefs about consequences**)
19. What do you think makes it most difficult to assess your treatment progress? (**Opportunity – Environmental context and resources; Opportunity – Social Influences**)
    a. **Probes:** lack of phone/computer, people who are close to you

**<u>If time permitting questions:</u>**

**Knowledge of patient portal and standardized questionnaires (PROMs)**

1. If you have used standardized questionnaires, will you share with me your personal experience with them?
    a. **Probes:** Did they impact the care you received?
2. If you use the patient portal (myhealth, Vanderbilt), how to you use it?
3. If you received a link on your phone to answer a couple questions, would you do it (why/why not)?

**1. Implementation of patient-reported outcome measures (PROMs) in routine clinical practice**

i) Do you have a sense of how these questionnaires affects your visit?
ii) If you could complete these questionnaires through the portal, would you?
iii) What about through text message?
iv) Do you have any thoughts about completing these questions?

**2. How these questionnaires might improve clinical care**

i) Have you discussed the results of the questionnaires with your doctor or other staff?
ii) Do you think this information could be useful?
    (1) Improve communication?
    (2) Monitor care between visits?
    (3) Examples?
iii) Do you think this information would be useful for certain people as opposed to others?

**3. Sharing PROM results with patients**

i) How would you feel about a clinician showing you results and talking them through with you?
ii) What are your thoughts about reviewing your own results through the patient portal?
iii) Is there any specific way you feel results should be displayed (e.g. portal, charts over time, etc.)?

**4. Patient suggestions**

a. Do you have any specific suggestions for us regarding these questionnaires? For example, are there other ways we could use these questionnaires to improve patient care?

**Closing questions:**

20. What do you think is the most important thing clinicians should know about supporting treatment between office visits?
21. Is there anything you think is important about supporting treatment for opioid use that I haven't asked?

**Demographics:**

May I ask, what is your Age: ________________
How would you identify yourself (gender, race, and ethnicity)?
Are you employed?
Do you have a place that you stay that you consider home?

Thank you again for your time to participate in this study. If you have any questions about the project, please contact Thomas Reese:.
